# High-risk *Escherichia coli* clones that cause neonatal meningitis and association with recrudescent infection

**DOI:** 10.1101/2023.10.05.23296362

**Authors:** Nguyen Thi Khanh Nhu, Minh-Duy Phan, Steven J. Hancock, Kate M. Peters, Laura Alvarez-Fraga, Brian M. Forde, Stacey B. Andersen, Thyl Miliya, Patrick N.A. Harris, Scott A. Beatson, Sanmarie Schlebusch, Haakon Bergh, Paul Turner, Annelie Brauner, Benita Westerlund-Wikström, Adam D. Irwin, Mark A. Schembri

**Author notes:** Wellcome-Wolfson Institute for Experimental Medicine, School of Medicine, Dentistry and Biomedical Sciences, Queen’s University Belfast, Belfast, United Kingdom (Steven J. Hancock). INRAE, Univ Montpellier, LBE, 102 Avenue des Etangs, Narbonne 11100, France. Corresponding authors: Professor Mark Schembri, Institute for Molecular Bioscience, The University of Queensland, Brisbane, Queensland 4072, Australia.; Dr Adam Irwin, Centre for Clinical Research, University of Queensland, Brisbane, Queensland, Australia.

## Abstract

Neonatal meningitis is a devastating disease associated with high mortality and neurological sequelae. *Escherichia coli* is the second most common cause of neonatal meningitis in full-term infants (herein NMEC) and the most common cause of meningitis in preterm neonates. Here we investigated the genomic relatedness of a collection of 58 NMEC isolates spanning 1974-2020 and isolated from seven different geographic regions. We show NMEC are comprised of diverse sequence types (STs), with ST95 (34.5%) and ST1193 (15.5%) the most common. No single virulence factor was conserved in all isolates; however, genes encoding fimbrial adhesins, iron acquisition systems, the K1 capsule, and O antigen types O18, O75 and O2 were most prevalent. Antibiotic resistance genes occurred infrequently in our collection. We also monitored the infection dynamics in three patients that suffered recrudescent invasive infection caused by the original infecting isolate despite appropriate antibiotic treatment based on antibiogram profile and resistance genotype. These patients exhibited severe gut dysbiosis. In one patient, the causative NMEC isolate was also detected in the fecal flora at the time of the second infection episode and after treatment. Thus, although antibiotics are the standard of care for NMEC treatment, our data suggests that failure to eliminate the causative NMEC that resides intestinally can lead to the existence of a refractory reservoir that may seed recrudescent infection.

## Introduction

Neonatal meningitis (NM) is a devastating disease with a mortality rate of 10-15% and severe neurological sequelae including hearing loss, reduced motor skills and impaired development in 30-50% of cases [1–3]. The incidence of disease is highest in low-income countries and occurs at a rate of 0.1-6.1/1000 live births [3]. *Escherichia coli* is the second most common cause of NM in full-term infants (herein NMEC), after group B *Streptococcus* (GBS) [4, 5], and the most common cause of meningitis in preterm neonates [5, 6]. Together, these two pathogens cause ∼60% of all cases, with on average one case of NMEC for every two cases of GBS [7, 8]. In several countries, NM incidence caused by GBS has declined due to maternal intrapartum antibiotic prophylaxis; however, NM incidence caused by *E. coli* remains the same [7, 9]. Moreover, NMEC is a significant cause of relapsed infections in neonates [10].

NMEC are categorised genetically based on multi-locus sequence type (ST) or by serotyping based on cell-surface O antigen (O), capsule (K) and flagella (H) antigens. Analysis of NMEC diversity in France revealed ∼25% of isolates belong to the ST95 clonal complex (STc95) [11], however, a global picture of NMEC epidemiology is lacking. NMEC possess a limited diversity of serotypes, dominated by O18:K1:H7, O1:K1, O7:K1, O16:K1, O83:K1 and O45:K1:H7, which together account for >70% of NMEC [12–15]. Notably, ∼80% of NMEC express the K1 capsule, a polysaccharide comprising linear homopolymers of α2-8-linked N-acetyl neuraminic acid [12, 16]. Apart from the K1 capsule, specific NMEC virulence factors are less-well defined, though studies have demonstrated a role for S fimbriae [17], the outer membrane protein OmpA [18], the endothelial invasin IbeA [19] and the cytotoxin necrotising factor CNF1 [20] in translocation of NMEC across the blood-brain barrier and pathogenesis. A large plasmid encoding colicin V (ColV), colicin Ia bacteriocins and several virulence genes including iron-chelating siderophore systems has also been strongly associated with NMEC virulence [21].

Despite being the second major NM aetiology, genomic studies on NMEC are lacking, with most reporting single NMEC complete genomes. Here we present the genomic analyses of a collection of 58 NMEC isolates obtained from seven different geographic regions over 46 years to understand virulence gene content, antibiotic resistance and genomic diversity. In addition, we provide a complete genome for 18 NMEC isolates representing different STs, serotypes and virulence gene profiles, thus more than tripling the number of available NMEC genomes that can be used as references in future studies. Three infants in our study suffered recrudescent invasive NMEC infection, and we show this was caused by the same isolate. We further revealed that patients that suffered recrudescent invasive infection had severe gut dysbiosis, and detected the infecting isolate in the intestinal microflora, suggesting NMEC colonisation of the gut provides a reservoir that can seed repeat infection.

## Results

### Establishment of an NMEC collection from geographically diverse locations

A collection of 52 NMEC isolates cultured from the blood or cerebrospinal fluid (CSF) of neonates with meningitis was established with the addition of six completely sequenced NMEC genomes available on the NCBI database. This yielded a final set of 58 NMEC isolates spanning 1974 to 2020 (Supplementary Table 1). The isolates were obtained from seven different geographic locations; Finland (n=17, 29.3%), Sweden (n=14, 24.1%), Australia (n=15, 25.9%), Cambodia (n=7, 12.1%), USA (n=3, 5.2%), France (n=1, 1.7%) and the Netherlands (n=1, 1.7%).

### ST95 and ST1193 are the two major STs of NMEC

Phylogenetic analysis was performed on the 58 NMEC isolates, with an additional eight well-characterised *E. coli* strains included for referencing (EC958, UTI89, MS7163, CFT073, UMN026, 536, APEC01, MG1655) (Supplementary Table 1). The NMEC isolates were diverse, and spanned phylogroups A, B2, C, D and F; the majority of isolates were from phylogroup B2 (n=48, 82.8%). Overall, the isolates belonged to 22 STs, of which 15 STs only contained one isolate. ST95 (n=20, 34.5%) and ST1193 (n=9, 15.5%) were the two most common NMEC STs (Figure 1, Supplementary Table 1). ST95 isolates were obtained over the entire study period, while ST1193 isolates were more recent and only obtained from 2013. Four isolates belonged to ST390 (6.9%), which is part of the STc95. One isolate belonged to a novel ST designated ST11637, which is part of the ST14 clonal complex (STc14) that also contains ST1193 (Figure 1, Supplementary Table 1). Isolates from other common phylogroup B2 extra-intestinal pathogenic *E. coli* (ExPEC) lineages, ST131, ST73 and ST69, as well as several STs associated with environmental sources such as ST48 and ST23, were detected in the collection. However, it is notable that the high incidence of NM associated with ST95 and ST1193 does not reflect the broader high prevalence of major ExPEC clones associated with human infections in the publicly available Enterobase database [22] (Supplementary Figure 1), suggesting ST95 and ST1193 exhibit specific virulence features associated with their capacity to cause NM.

**Figure 1.**
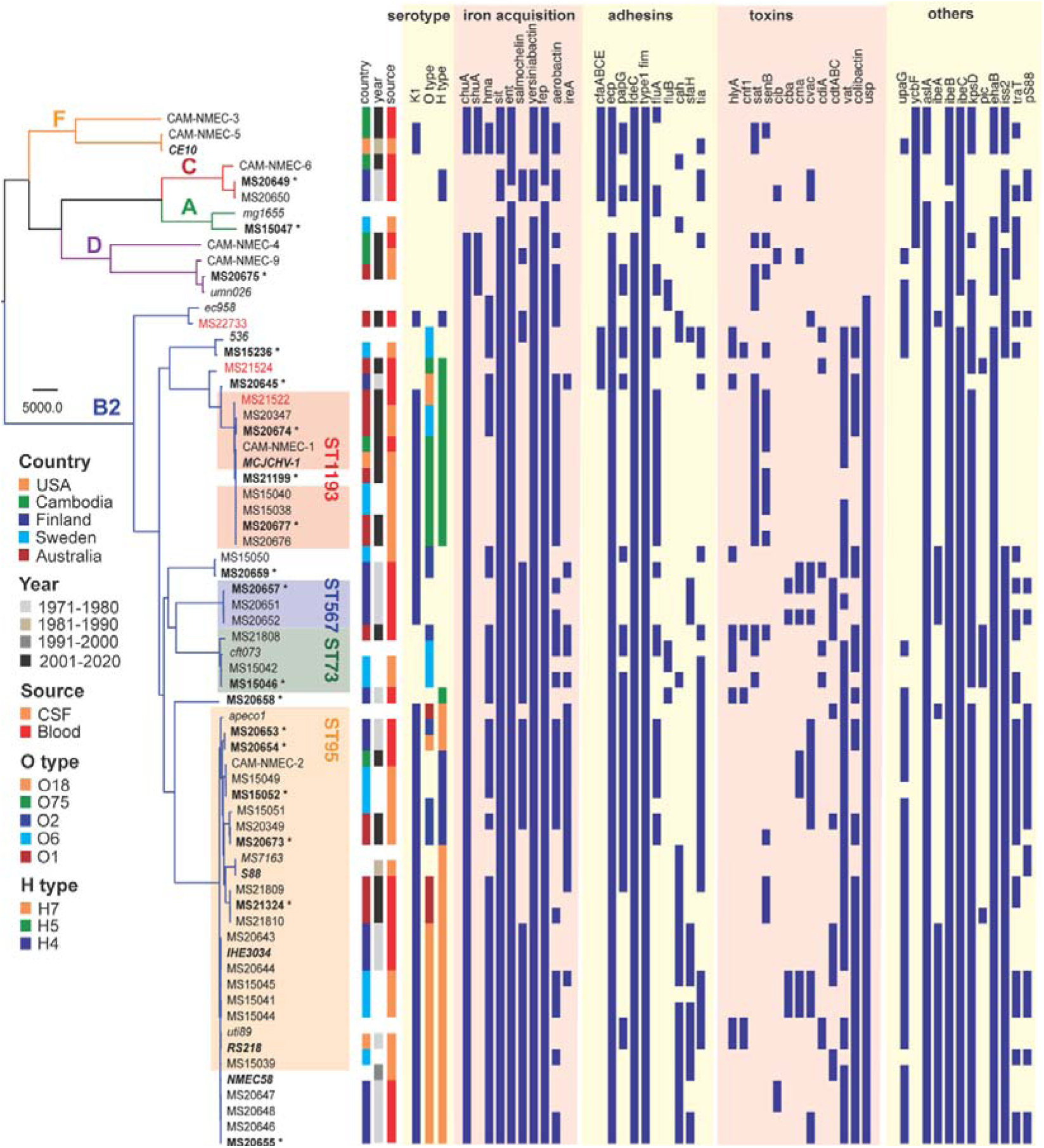
Maximum likelihood phylogram displaying the relationship of the NMEC isolates with their associated serotype and virulence factor profile. Non-NMEC isolates used in the analysis for referencing are italicized. The phylogram was built and recombination regions removed employing Parsnp, using 185,911 core single nucleotide polymorphisms (SNPs) and NMEC strain IHE3034 as the reference. The scale bar indicates branch lengths in numbers of SNPs. NMEC isolates with available complete genomes are bold-italicized, while NMEC isolates that were completely sequenced in this study are indicated in bold and marked with an asterisk. The NMEC isolates that caused recrudescent invasive infection in this study are indicated in red. Branches are colored according to phylogroups: orange, phylogroup F; red, phylogroup C; green, phylogroup A; violet, phylogroup D and blue, phylogroup B2. The presence of specific virulence factors is indicated in dark blue. The phylogeny can be viewed interactively at http://microreact.org/project/oNfA4v16h3tQbqREoYtCXj-high-risk-escherichia-coli-clones-that-cause-neonatal-meningitis-and-association-with-recrudescent-infection.

Eighteen NMEC isolates were completely sequenced using complementary long-read Oxford Nanopore Technology, enabling accurate comparison of NMEC genome size, genomic island composition and location, and plasmid and prophage diversity (Supplementary Table 2). These isolates spanned the diversity in the collection, representing 11 different STs, including two ST1193 isolates (one with the dominant O75:H5 serotype and one with an unusual O6:H5 serotype), five ST95 isolates with different serotypes, and one isolate from the novel ST11637.

### Antibiotic resistance in NMEC

Antibiotic resistance profiling revealed an overall low level of resistance in the collection. The ST1193 isolates contained fluoroquinolone resistance defining mutations in *gyrA* (S83L D87N) and *parC* (S80I), as previously described for this lineage [23]. In addition, 77.8% of ST1193 isolates (7/9 isolates) also harboured at least one gene conferring resistance to aminoglycosides (*aac(3)-IId*, *aadA5*, *aph(3’’)-Ib* and *aph(6)-Id*), trimethoprim (*dftA17*) and sulphonamides (*sul1* and *sul2*) (Supplementary Figure 2). Six out of the seven isolates from Cambodia had more than one antibiotic resistance gene, likely reflecting increased antibiotic resistance rates in this region [24]. Indeed, in addition to *gyrA* and *parC* mutations for fluoroquinolone resistance, CAM-NMEC-6 contains 14 antibiotic resistance genes (including resistance to third-generation cephalosporins and carbapenems) and CAM-NMEC-4 contains 11 antibiotic resistance genes (Supplementary Figure 2).

**Figure 2.**
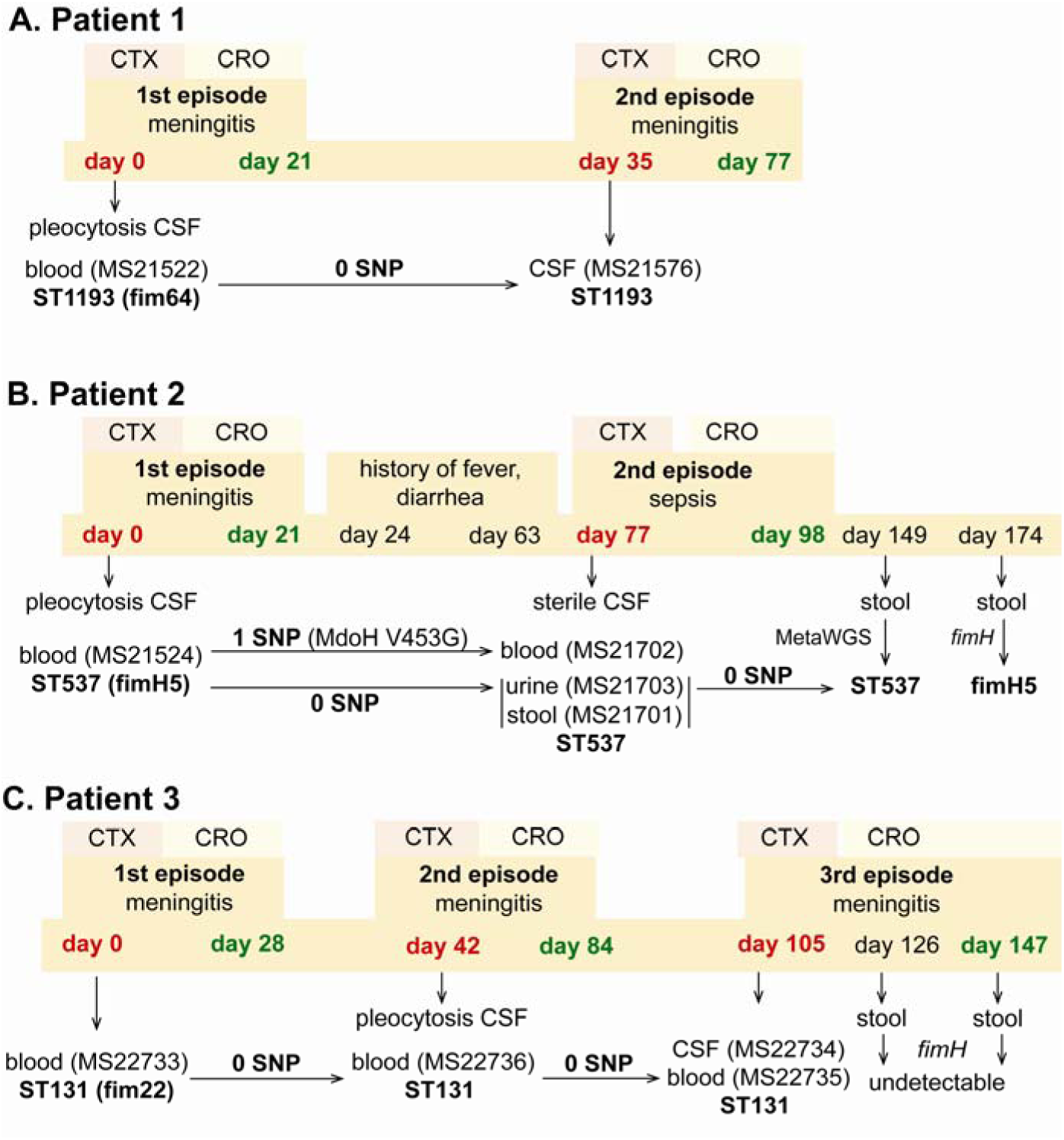
Infection and treatment profile of patients suffering NM and recrudescent invasive infection. Indicated is the hospital admission history of patients, together with the timeline of sample collection, identified *E. coli* isolates and their infection source, and isolate identification based on whole genome sequencing, metagenomic sequencing (MetaWGS) or *fimH* amplicon sequencing. Genomic relatedness is indicated based on the number of single nucleotide polymorphisms (SNPs). The time of admission for the initial episode is indicated as Day 0, with subsequent timepoints indicated as days post initial admission. Admission and discharge days are indicated in red and green, respectively.

### Virulence factors in NMEC

The isolates exhibited variable distribution of virulence genes previously linked to NMEC pathogenesis. The most prevalent genes were those involved in iron uptake, including the enterobactin (98%), yersiniabactin (98%), aerobactin (62%) and salmochelin (55%) siderophore systems, and the heme receptors *chuA* (93%) and *hma* (62%) (Figure 1). Also common were the *sitABCD* genes encoding an iron/manganese transporter (98%). The presence of fimbrial and afimbrial adhesins was also diverse. The most prevalent adhesins were type 1 fimbriae (100%), *mat* (*ecp*) fimbriae (98%) and the *fdeC* adhesin (98%). Genes encoding P and S fimbriae were detected in 36% and 22% of NMEC isolates, respectively. The most prevalent toxin was the uropathogenic-specific genotoxin *usp* (83%), which was only found in phylogroup B2 isolates. Other toxin genes encoding the serine protease autotransporters Vat (65% prevalence) and Sat (29%), hemolysin (12%) and cytotoxic necrotizing factor-1 (7%) were less prevalent. Additional virulence genes included the *aslA* arylsulfatase (95%), the *iss* lipoprotein (76%) and the *ibeA* invasin (33%). The ColV-plasmid was present in 33% of the isolates (Figure 1, Supplementary Table 1). Direct comparison of virulence factors between ST95 and ST1193, the two most dominant NMEC STs, revealed that the ST95 isolates (n = 20) contained significantly more virulence factors than the ST1193 isolates (n = 9); *P*-value < 0.001, Mann-Whitney two-tailed unpaired test (Supplementary Table 1, Supplementary Figure 3).

**Figure 3.**
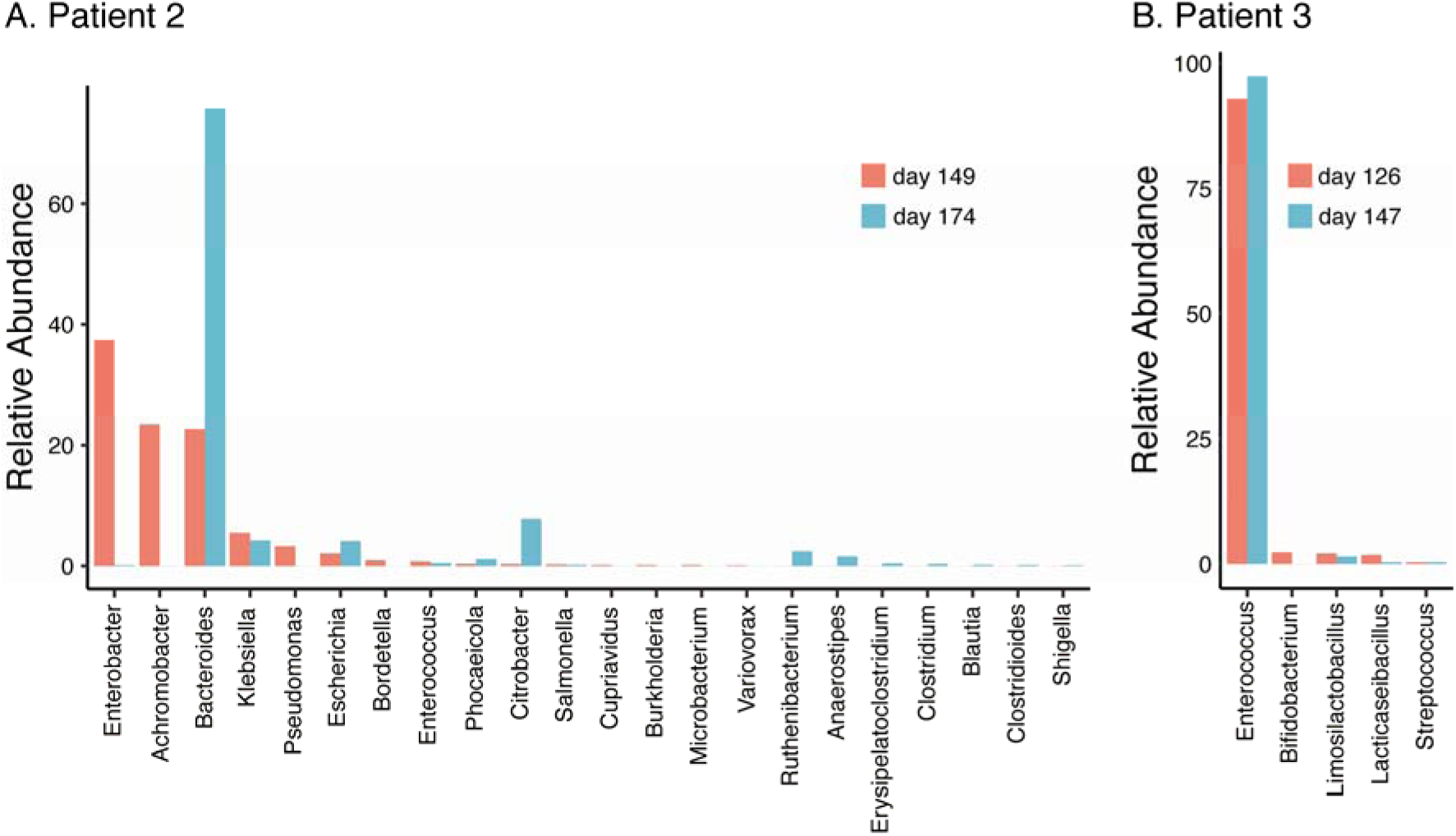
Relative abundance of bacterial genera (≥0.01%) in the gut microbiome of patient 2 at 8- and 12-week follow-up post relapsed infection (days 149 and 174 after initial admission) (A) and patient 3 during treatment and at discharge after the third episode (days 126 and 147 after initial admission) (B).

### NMEC comprise a dominant K1 capsule type and a limited pool of O and H serotypes

The capsule type of the NMEC isolates was determined by *in silico* typing. K1 was the dominant capsule type in the collection (43/58 isolates, 74.1%) (Figure 1). Thirty-four of these isolates were available for capsule testing, and we confirmed K1 expression by ELISA in all but two isolates (Supplementary Figure 4). Other capsule types included K2, K5 and K14 (Supplementary Table 1). A capsule type could not be resolved for 12 isolates, of which eight did not possess a Group II or Group III capsule type based on the absence of the conserved *kpsD* gene (Figure 1, Supplementary Table 1).

**Figure 4.**
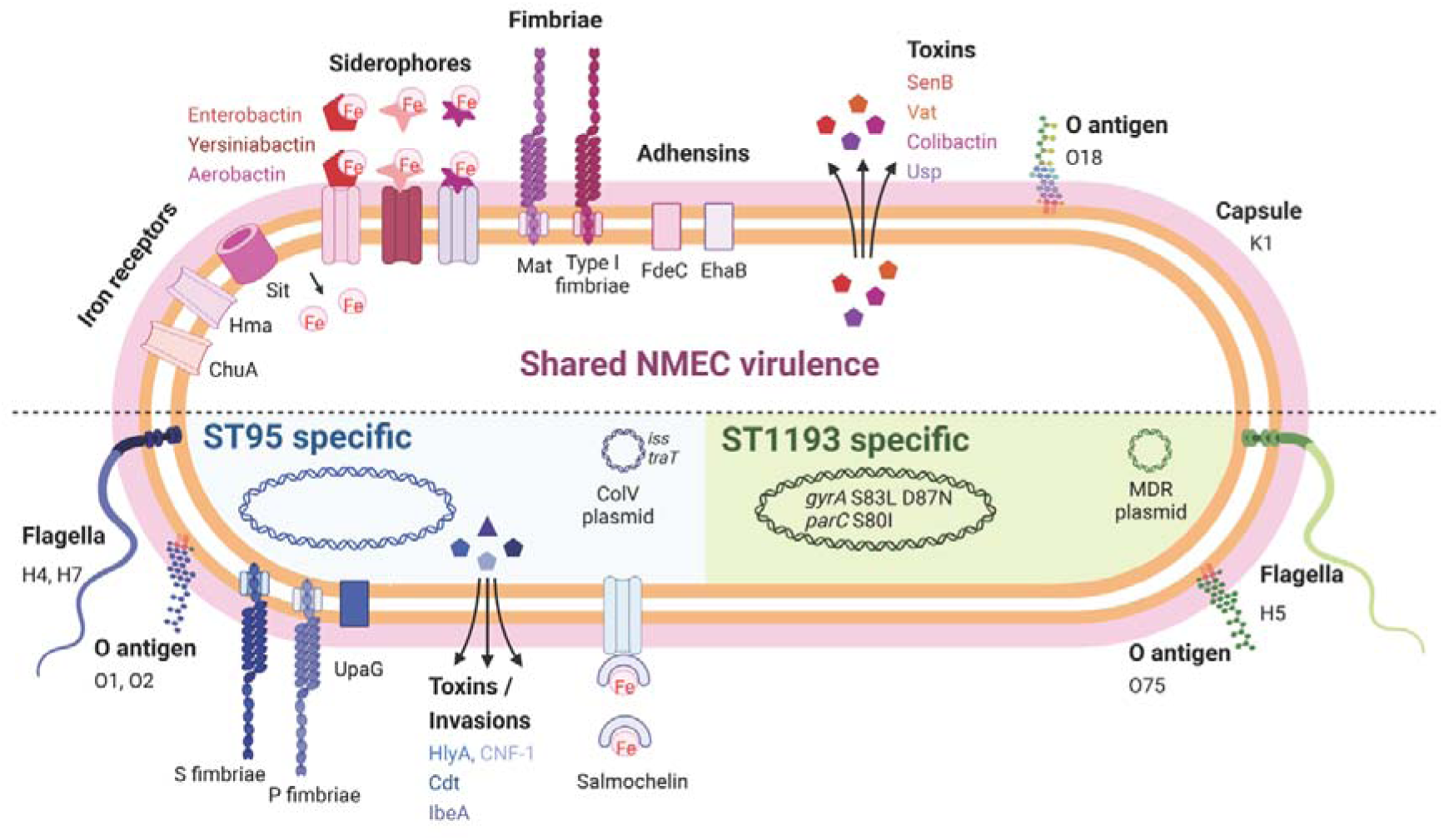
Summary of key NMEC virulence genes based on genome profiling performed in this study. Shown are shared virulence genes common to most NMEC, as well as ST95- and ST1193-specific NMEC virulence genes. Figure created with BioRender.com.

*In silico* O-antigen (O) and flagella (H) serotypes were also determined. O18 was the most common O type (n=16, 27.6%), followed by O75 (n=8, 13.9%) and O2 (n=7, 12.1%). The most dominant H types were H7 (n=19, 32.8%), H5 (n=13, 22.4%) and H4 (n=9, 15.5%). The most common serotype was O18:H7:K1 (n=14, 24.1%); these isolates belonged to the STc95 (nine ST95, four ST390 and one ST416). The second most common serotype was O75:H5:K1 (n=8, 13.8%); six isolates from ST1193 possessed this serotype.

### NMEC can cause recrudescent invasive infection despite appropriate antibiotic treatment

During 2019 - 2020, three patients from which NMEC isolates were originally cultured suffered recrudescent invasive infection (Figure 1; MS21522, MS21524 and MS22733), providing an opportunity to compare the infecting isolates over time using whole genome sequencing. In all cases, the infecting *E. coli* isolates were susceptible to the therapy, which comprised cefotaxime (50mg/kg/dose 8 hourly), switched to ceftriaxone (100mg/kg/day) to facilitate home parenteral antibiotic administration. Bacterial culture was performed from blood, CSF, urine and/or stool during the infection period (Figure 2). These patients were from different regions in Australia.

#### Patient 1

Patient 1 (0-8 weeks of age) was admitted to the emergency department with fever, respiratory distress and sepsis. The child was diagnosed with meningitis based on a cerebrospinal fluid (CSF) pleocytosis (>2000 white blood cells; WBCs, low glucose, elevated protein), positive CSF *E. coli* PCR and a positive blood culture for *E. coli* (MS21522). Two weeks after completion of a 3-week course of appropriately dosed therapy with third-generation cephalosporins as described above, the child developed similar symptoms of fever and irritability. Lumbar puncture was performed and the CSF culture was positive for *E. coli* (MS21576). Both the initial blood culture isolate and the relapse CSF isolate were non-susceptible to ciprofloxacin and gentamycin, and whole genome sequencing revealed they were identical (ST1193 O18:K1:H5; *fimH*64), with no single nucleotide polymorphisms (SNPs) nor indels (Figure 2A). Unlike the typical ST1193 O75 serotype [23], this isolate contained a unique O18 serotype. The isolate possessed mutations in *gyrA* (S83L D87N) and *parC* (S80I), which explain its resistance to ciprofloxacin, as well as a multidrug resistance IncF plasmid containing genes conferring resistance to aminoglycosides (*aac(3)-IId*, *aadA5*, *aph(3’’)-Ib* and *aph(6)-Id*), trimethoprim (*dfrA17*), sulphonamides (*sul1* and *sul2*) and macrolides (*mphA*) (Supplementary Figure 2). Treatment of the relapse was extended to six weeks of intravenous therapy. At follow-up, no anatomical or immunological abnormality was identified and development is normal.

#### Patient 2

Patient 2 (0-8 weeks of age) presented to the emergency department with a febrile illness. Blood and urine cultures on admission were positive for *E. coli*. CSF taken 24 hours after treatment revealed pleocytosis (>300 WBCs, >95% polymorphs) but no bacteria were cultured. The patient completed a 3-week course of appropriately dosed antibiotic therapy with third-generation cephalosporins. In the 6-week period after discharge, the child had several short admissions to hospital, but no infection was identified. At 11 weeks post initial infection, the child was re-admitted to hospital with high fever. CSF cultures were negative and microscopy was normal, but cultures from blood, urine and faeces were all positive for *E. coli*. Whole genome sequencing revealed that all isolates belonged to ST537 O75:H5 (*fimH*5; STc14). Pairwise comparison of the recrudescent isolates showed that the urine and fecal isolates were identical to the original isolate, while the blood isolate contained one nonsynonymous SNP in the *mdoH* gene encoding a glucan biosynthesis glucosyltransferase (T1358G; V453G). This mutation is located in the large cytoplasmic domain of MdoH likely involved in polymerisation of glucose from UDP glucose; the isolate exhibited a mucoid colony morphology suggestive of increased colanic acid production. The isolates did not possess plasmids nor antibiotic resistance genes. The infant experienced recurrent urinary tract infections with *E. coli* and other urinary pathogens through infancy despite normal urinary tract anatomy. At follow-up, no other history of invasive infection nor identified immunodeficiency were noted, and the child was reported to be developing normally.

#### Patient 3

Patient 3 (0-8 weeks of age) was admitted to the paediatric intensive care unit with fever and seizures. CSF and blood cultured a fully susceptible *E. coli*. Two weeks after completing a four-week course of appropriate therapy with third-generation cephalosporins, the infant was readmitted to hospital with fever and irritability, with further investigation identifying *E. coli* in CSF, urine and blood. Three weeks after the completion of the six-week treatment course, the infant experienced a second relapse, with *E. coli* isolated from both CSF and blood. Whole genome sequencing revealed that all isolates were identical and belonged to ST131 O25b:K1:H4 (*fimH*22). These isolates contained a ColV-virulence plasmid, but did not harbour acquired antibiotic resistance genes. The infant received a further 6-week course of therapy. Extensive imaging studies including repeated MRI imaging of the head and spine, CT imaging of the head and chest, ultrasound imaging of abdomen and pelvis, and nuclear medicine imaging did not show a congenital malformation or abscess. Immunological work-up did not show a known primary immunodeficiency. At follow-up, speech delay is reported but no other developmental abnormality.

### The gut as a reservoir to seed recrudescent infection

In all three patients that suffered NM and recrudescent invasive infection, the causative isolates were susceptible to third-generation cephalosporins, suggesting the existence of a persistent reservoir that could evade the cidal effect of antibiotic treatment and seed repeat infection. Indeed, the fact that the causative *E. coli* isolate was detected from a fecal sample at the time of the recrudescent infection in patient 2 (day 77 after initial admission), suggests that NMEC could persist in the gut and cause repeat infection, an observation that has also been reported for uropathogenic *E. coli* that cause recurrent urinary tract infection [25] and acute pyelonephritis in infants [26]. Therefore, we retrospectively examined available stored fecal samples from patient 2 at 8- and 12-week follow-up visits post recrudescent infection (days 149 and 174 after initial admission) and patient 3 during treatment and at discharge after the third episode (days 126 and 147 after initial admission) using short-read metagenomic sequencing (Figure 3). Although no fecal samples were available for comparative analysis from either patient prior to antibiotic treatment, we observed a low level of diversity in the composition of the microbiome of both patients, consistent with severe dysbiosis. The microbiome of patient 2 was dominated by *Enterobacter* (37.4% relative abundance), *Achromobacter* (23.4% relative abundance) and *Bacteroides* (22.7% relative abundance) genera at 8-weeks post recrudescent infection (day 149 after initial admission), and by *Bacteroides* genera (75.8% relative abundance) at 12-weeks post recrudescent infection (day 174 after initial admission). The relative abundance of *E. coli* was 2.05% and 4.1% in each of these samples, respectively, and further analysis using StrainGE [27] showed that the isolates were most closely matched to the original causative MS21524 isolate. We further employed complementary long-read metagenomic sequencing to analyse the 8-week post relapse infection sample, which enabled construction of a complete *E. coli* genome that was identical to the causative ST537 (*fimH*5) isolate (Figure 2, Figure 3; Supplementary Table 3). In the 12-week post recrudescent infection fecal sample from patient 2, amplicon sequencing targeting *fimH* identified the presence of *E. coli* with the same *fimH* type as the causative isolate (*fimH5*). Thus, two independent analyses of samples taken 4 weeks apart demonstrated the existence of the *E. coli* ST537 isolate in the intestinal microflora of patient 2. In patient 3, the microbiome was dominated by *Enterococcus* genera at both time points examined (93% and 97.4% relative abundance). We were unable to detect *E. coli* by *fimH* amplicon sequencing and the relative abundance of *E. coli* in these fecal samples was extremely low (<0.01%) based on metagenomic sequencing (Supplementary Table 3). The extensive dysbiosis revealed in this patient is likely an outcome of the three rounds of antibiotic treatment.

## Discussion

In this study, we present a genomic analysis of 58 NMEC isolates obtained over 46 years spanning seven different geographic locations and reveal a dominance of ST95 and ST1193. We also provide direct evidence to implicate the gut as a reservoir for recrudescent invasive infection in some patients despite appropriate antibiotic treatment.

The majority of the NMEC isolates in our study belonged to phylogroup B2 (82.8%), an observation consistent with other reports [28, 29]. These isolates were predominantly from two major STs, ST95 and ST1193. ST95 represents a major clonal lineage responsible for urinary tract and bloodstream infections [30, 31], and were identified throughout the period of investigation. This lineage was also previously shown to cause ∼25% of NM cases in France in the period 2004-2015 [11], demonstrating its enhanced capacity to cause disseminated infection in newborns. ST1193, on the other hand, was first identified in 2012 [32], and is the second most common fluoroquinolone-resistant *E. coli* lineage after ST131 [23, 33]. ST1193 causing NM was first reported in the USA in 2016 [34]. Here, ST1193 accounted for 15.5% of NMEC isolates, all of which were obtained from 2013, and was the dominant lineage since this time. This is consistent with a report in China that showed ST1193 was the most common NMEC (21.4%), followed by ST95 (17.9%), between 2009-2015 [35]. Concerningly, the ST1193 isolates examined here carry genes encoding several aminoglycoside-modifying enzymes, generating a resistance profile that may lead to the clinical failure of empiric regimens such as ampicillin and gentamicin, a therapeutic combination used in many settings to treat NM and early-onset sepsis [36, 37]. This, in combination with reports of co-resistance to third-generation cephalosporins for some ST1193 isolates [23, 35], would limit the choice of antibiotic treatments. The dominance of both ST95 and ST1193 in our collection is notable since other widespread *E. coli* phylogroup B2 lineages such as ST131, ST73, ST69 and ST12 do not cause similar rates of NM disease. We speculate this is due to the prevailing K1 polysialic acid capsule serotype found in ST95 and the newly emerged ST1193 clone [23, 38] in combination with other virulence factors [15, 28, 29] (Figure 4) and the immature immune system of preterm infants. Understanding the risk of these clones, as well as perinatal transmission and antibiotic resistance patterns, may inform the appropriateness of interventions such as maternal screening or antimicrobial prophylaxis.

Although reported rarely, recrudescent invasive *E. coli* infection in NM patients, including several infants born pre-term, has been documented in single study reports [39, 40]. In these reports, infants received appropriate antibiotic treatment based on antibiogram profiling and no clear clinical risk factors to explain recrudescence were identified, highlighting our limited understanding of NM aetiology. Here, we tracked NMEC recurrence using whole genome sequencing in three patients that suffered NM and recrudescent invasive infection, and demonstrated that the isolate causing recrudescence was the same as the original causative isolate and susceptible to the initial antibiotic therapy. In one patient (patient 2), we identified the causative isolate in the stool at days 77, 149 and 174 after initial detection in the bloodstream, providing direct evidence of persistence in the gut, and implicating this site as a reservoir to seed repeat infection. This isolate belonged to ST537 (serotype O75:H5) and is from the same clonal complex as ST1193 (i.e. STc14).

This study had several limitations. First, our NMEC collection was restricted to seven geographic regions, a reflection of the difficulty in acquiring isolates causing this disease. Second, we did not have access to a complete set of stool samples spanning pre- and post-treatment in the patients that suffered NM and recrudescent invasive infection. This impacted our capacity to monitor *E. coli* persistence and evaluate the effect of antibiotic treatment on changes in the microbiome over time. Third, we did not have access to urine or stool samples from the mother of the infants that suffered recrudescence, and thus cannot rule out mother-to-child transmission as a mechanism of reinfection. Finally, we did not have clinical data on the weeks of gestation for all patients, and thus could not compare virulence factors from NMEC isolated from preterm versus term infants. Regardless, our study describes the genomic diversity of NMEC, highlighting ST95 and ST1193 as the most important clonal lineages associated with this devastating disease. Although antibiotics are the standard of care for NMEC treatment, we show that even when appropriate antibiotics are used, in some cases they do not eliminate the causative NMEC that resides intestinally. Together with associated antibiotic-driven dysbiosis, this reveals a need to consider diagnostic and therapeutic interventions to mitigate the risk of recrudescent infection.

## Methods

### Ethics statement

The study received ethical approval from the Children’s Health Queensland Human Research Ethics Committee (LNR/18/QCHQ/45045). Precise patient details have been removed for ethics compliance; additional de-identified details are available from the corresponding author upon request.

### Bacterial isolates

A collection of 52 NMEC isolates obtained from 1974 to 2020 was achieved from Sweden, Finland, Cambodia and Australia. Isolates were stored in glycerol at -80°C until use. All isolates were cultured in Lysogeny broth. The collection comprised 42 isolates from confirmed meningitis cases (29 cultured from CSF and 13 cultured from blood) and 10 isolates from clinically diagnosed meningitis cases (all cultured from blood) (Table S1). This collection was complemented by the addition of six completely sequenced NMEC genomes available on the NCBI database, namely strains IHE3034, RS218, S88, NMEC58, MCJCHV-1 and CE10.

### DNA extraction, genome sequencing and analyses

Genome sequencing was performed using paired-end Illumina methodology. Illumina sequencing data were processed by removing adapters and low-quality reads using Trimmomatic v0.36 [41], with a minimum quality score of 10 and minimum read length of 50. Trimmed reads were *de nov*o assembled using SPAdes v3.12.0 [42] with default parameters. Draft assemblies of the 52 NMEC isolates from this study, together with six complete NMEC genomes and eight complete genomes from other characterised *E. coli* representing different phylogroups, were subjected to phylogenetic analysis using parsnp v1.5.3 [43]. A subset of 18 isolates were additionally sequenced using Oxford Nanopore Technology long-read sequencing (Nanopore). Complete NMEC genomes were achieved using a combination of Illumina short-read and Nanopore long-read data and analysis employing the MicroPIPE tool [44].

### *In silico* and molecular analyses

Virulence-associated genes, antibiotic resistance genes, plasmids and serotyping were evaluated using ABRicate (https://github.com/tseemann/abricate) with built-in databases [45–48], with the percentage nucleotide identity and coverage cut-off set at 90% and 80%, respectively. Capsule typing was performed employing Kaptive [49] using an in-house *E. coli* capsule database [38] and manually checked. Chromosomal point mutations associated with antibiotic resistance were detected using PointFinder [50]. FimH amplicon sequencing was performed as previously described [51, 52]; allelic variants were identified using FimTyper [53].

### K1 ELISA

K1 capsule expression was detected by ELISA using an anti-polysialic acid antibody single chain Fv fragment [54] as the primary antibody, anti-His antibody and alkaline phosphatase anti-mouse IgG as the secondary and tertiary antibodies, respectively; p-nitrophenylphosphate (Sigma) was used as the substrate. Optical density was measured at 420 nm.

### Metagenomic sequencing and analyses

Metagenomic sequencing was performed on DNA extracted from fecal samples using the Illumnina NovaSeq6000 platform. Adapters and low-quality reads were trimmed using Trimmomatic v0.36 [41], employing a minimum quality score of 10 and minimum read length of 50. Sequencing reads corresponding to human DNA were discarded by mapping the trimmed reads to the human genome hg38 (accession number GCA_000001405.29) using bowtie2 [55]. Taxonomical profiling was performed with Kraken2 [56] followed by Bracken [57].

### Long-read metagenomic sequencing

Long-read metagenomic sequencing was performed on DNA extracted from a fecal sample. A HiFi gDNA library was prepared using the SMRTbell Express Template Prep Kit 2.0 (PacBio, 100-938-900) according to the low input protocol (PacBio, PN 101-730-400 Version 06 [June 2020]). As the sample DNA was already fragmented with a tight peak (mode size 9.4 kb), no shearing was performed; the sample was concentrated using Ampure PB beads (PacBio, PCB-100-265-900) and used directly as input into library preparation. The entire quantity of purified DNA (360ng) was used to make the library as follows. The DNA was treated to remove single-stranded overhangs, followed by a DNA damage repair reaction and an end-repair/A-tailing reaction. Overhang barcoded adapters were ligated to the A-tailed library fragments, followed by a nuclease treatment to remove damaged library fragments, and then purification with AMPure PB beads. The library was size-selected to remove fragments <3kb using AMPure PB beads. The final purified, size-selected library was quantified on the Qubit fluorometer using the Qubit dsDNA HS assay kit (Invitrogen, Q32854) to assess concentration, and run on the Agilent Femto Pulse using the 55 kb BAC Analysis Kit (Agilent, FP-1003-0275) to assess fragment size distribution.

Sequencing was performed using the PacBio Sequel II (software/chemistry v10.1). The library pool was prepared for sequencing according to the SMRT Link (v10.1) sample setup calculator, following the standard protocol for Diffusion loading with Ampure PB bead purification, using Sequencing Primer v5, Sequel II Binding Kit v2.2 and the Sequel II DNA Internal Control v1. Adaptive loading was utilised, with nominated on-plate loading concentration of 80pM. The polymerase-bound library was sequenced on 1 SMRT Cell with a 30-hour movie time plus a 2-hour pre-extension using the Sequel II Sequencing 2.0 Kit (PacBio, 101-820-200) and SMRT Cell 8M (PacBio, 101-389-001).

After sequencing, the data was processed to generate CCS reads and demultiplex samples using the default settings of the CCS with Demultiplexing application in SMRT Link (v10.1). The demultiplexed reads were assembled *de novo* using Hifiasm [58]. Assembled contigs were subject to taxonomic profiling using kraken2 [56] and fastANI [59]. Contigs taxonomically assigned as *Escherichia coli* were subjected to *in silico* sequence typing using MLST (https://github.com/tseemann/mlst) and mlst profiles from PubMLST [60].

### Author Contributions

Conceptualization, MAS, AB, BW-W and ADI.; Investigation, NTKN, M-DP, SJH, KMP, LA-F, BMF, SBA, and TM.; Formal analysis, NTKN, M-DP, ADI and MAS., Resources, PNAH, SAB, SS, HB, PT, AB, BW-W, ADI and MAS.; Writing – Original Draft, NTKN, ADI and MAS.; Writing – Review & Editing, All Authors.; Supervision, ADI and MAS.; Funding Acquisition, NTKN, M-DP, PNAH, SAB, SS, PT, ADI, MAS.

### Data sharing

Genome sequence data have been deposited in the Sequence Read Archive under the BioProjects PRJNA757133 and PRJNA893826. Sample accession numbers are listed in Supplementary Table 4.

## Data Availability

All data produced in the present work are contained in the manuscript

## Acknowledgments

The authors would like to thank Michelle Bauer for technical expertise and the laboratories contributing the isolates, Pathology Queensland and Mater Pathology. At the time of the study SS was affiliated with Mater Pathology, South Brisbane, Australia. This work was supported by project grants APP1181958 and APP2001431 (to MAS, MDP and NTKN) and an Investigator grant GNT1197743 (to ADI) from the National Health and Medical Research Council of Australia (NHMRC), a Children’s Hospital Foundation Innovator grant (50270, to MAS, ADI, PNAH, SAB and SS), an Australian Infectious Diseases Research Centre grant (to MAS, ADI and NTKN), a grant from the Genome Innovation Hub at the University of Queensland, and a grant from the Wellcome Trust to PT (Grant number 220211). For the purpose of open access, the author (PT) has applied a CC BY public copyright licence to any Author Accepted Manuscript version arising from this submission.

## Supplementary Figure Legends

**Figure S1.**
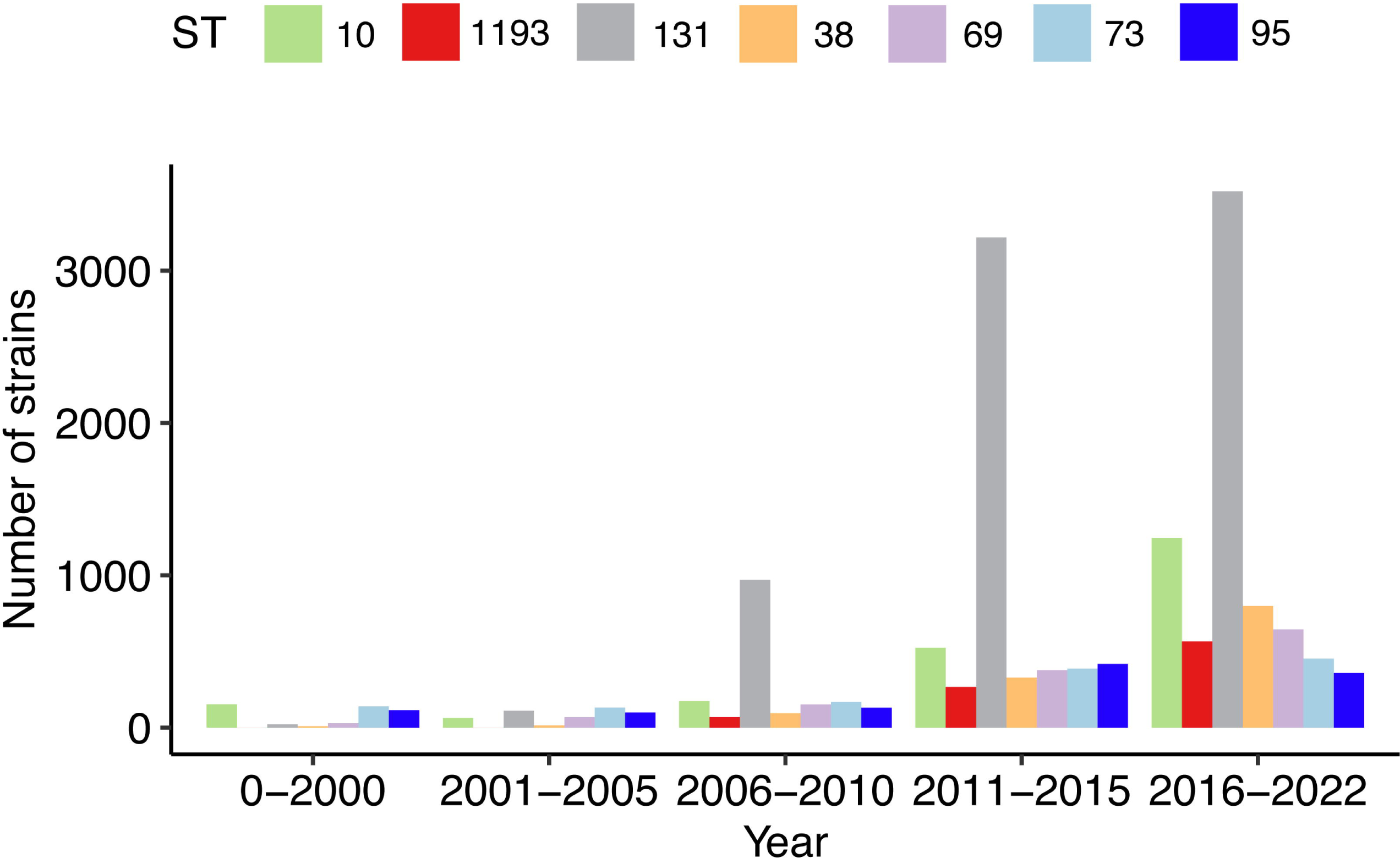
Number of human-derived *E. coli* strains from ST95, ST1193, ST38, ST131, ST73, ST10 and ST69 available in the Enterobase database. Strains were stratified based on their year of isolation, spanning the periods before 2000, 2001-2005, 2006-2010, 2011-2015 and 2016-2022.

**Figure S2.**
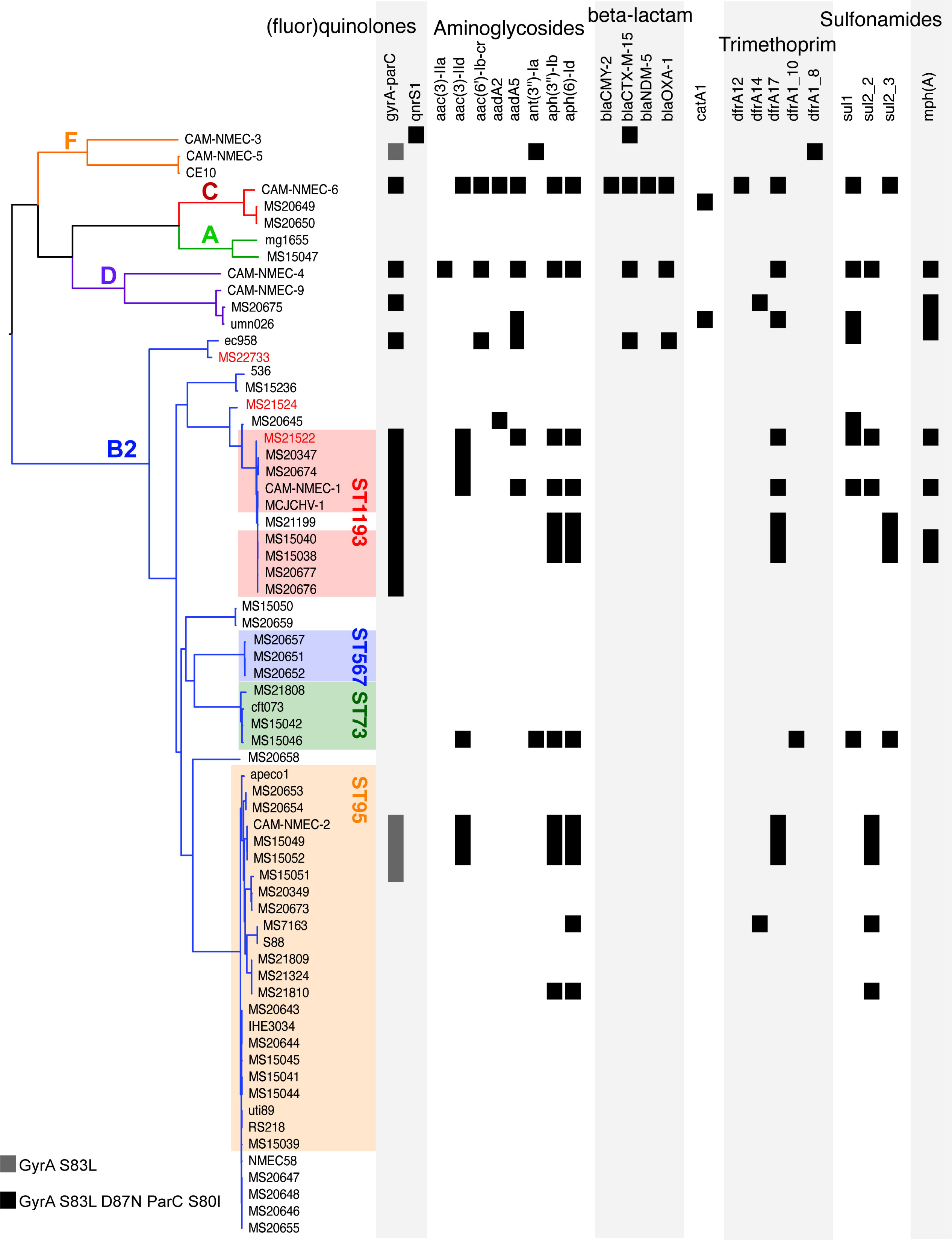
Antibiotic resistance gene profile of NMEC strains in the collection. The presence of each resistance gene is denoted by black shading.

**Figure S3.**
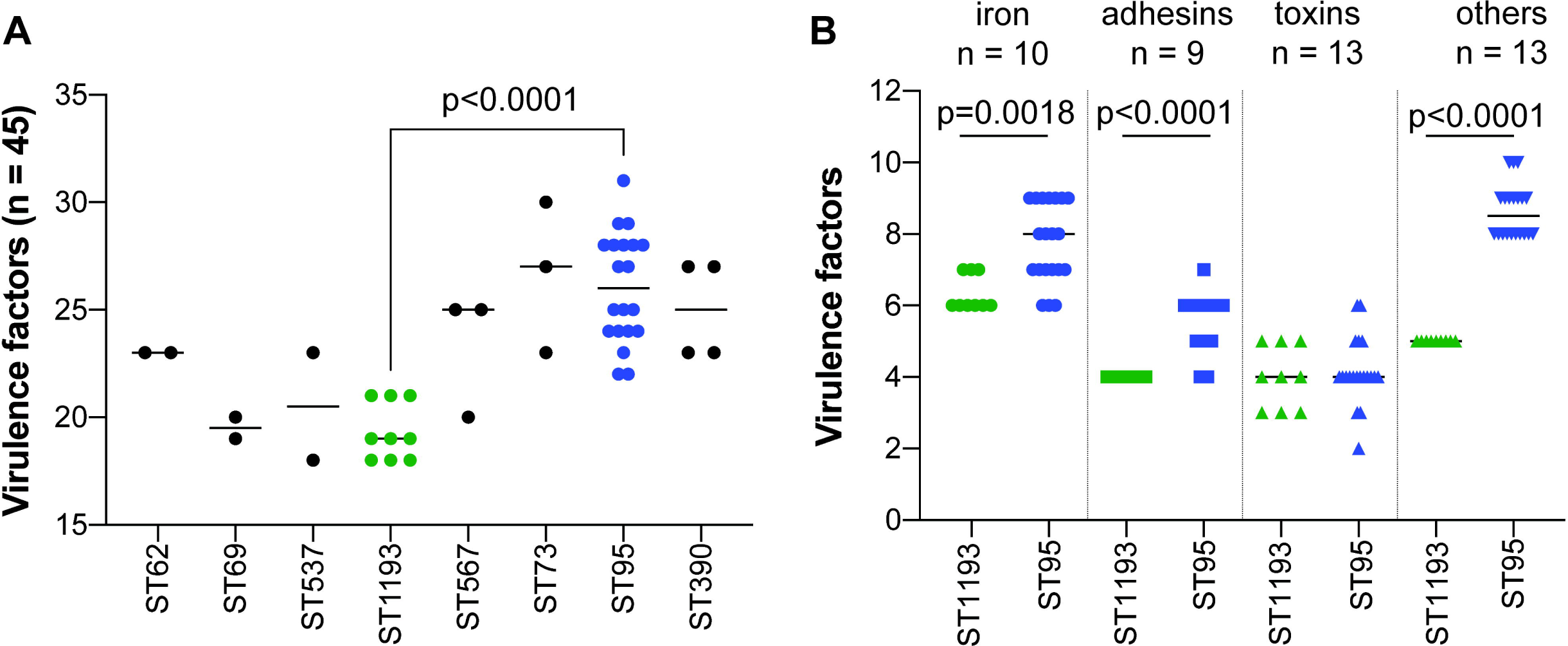
ST95 NMEC strains contain more virulence factors than ST1193 NMEC strains. (A) The number of virulence genes (grouped as in Fig.1) for each strain within each ST. (B) The number of virulence genes grouped by their functions in ST95 versus ST1193 strains. P-value were calculated using Mann-Whitney two-tailed unpaired test.

**Figure S4.**
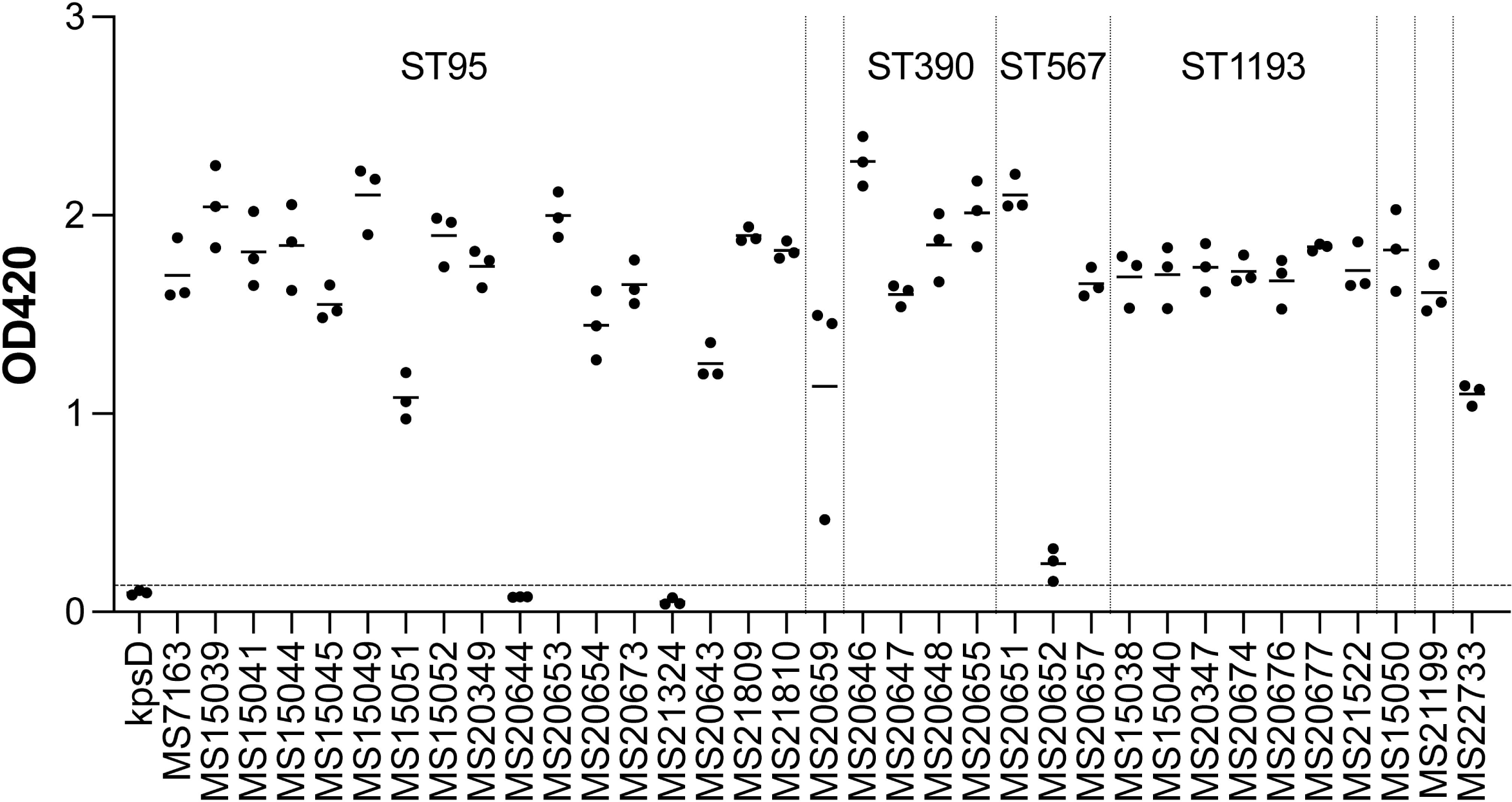
K1 capsule production in NMEC. K1 capsule production was detected by ELISA using a monoclonal antibody specific for polysialic acid. Strains with an OD_420_ > 0.133 (mean + 3 standard deviations of a negative control *kpsD* mutant; dashed line) were considered positive for K1 capsule production. Data points represent independent biological replicates with horizontal lines as the mean.

## Supplementary Tables

**Supplementary Table 1.** Isolates used in this study.

**Supplementary Table 2.** Completely sequenced NMEC isolates.

**Supplementary Table 3.** Metagenomic sequence analysis.

**Supplementary Table 4.** Accession numbers of strains sequenced in the study

